# Prenatal therapies to improve outcomes in gastroschisis: A systematic scoping review protocol

**DOI:** 10.1101/2025.06.07.25329164

**Authors:** MF Varela, J Reed, M Oria, S Kosaka, E Lopriore, JL Peiro

## Abstract

**Introduction:** Gastroschisis is a congenital birth defect with a rising incidence. The herniation of intestines into the amniotic cavity leads to prenatal bowel injury, which contributes to significant postnatal morbidity and mortality. Current management focuses on postnatal interventions, though bowel injury begins in utero. Emerging interest in prenatal therapies aims to mitigate this injury and improve outcomes. However, none of these interventions have been widely adopted in clinical practice, highlighting the need for further research and validation.

**Objectives:** Outline the procedures to conduct a systematic scoping review of prenatal interventions to improve outcomes for gastroschisis, identifying gaps in the literature to guide future research and inform clinical practice.

**Methods and analysis:** This systematic scoping review will follow the PRISMA-ScR guidelines. We will include studies on prenatal interventions for gastroschisis in both human and animal models. Key databases including MEDLINE, EMBASE, Scopus, and others will be searched. Gray literature and clinical trial registries will also be reviewed. Studies will be screened, selected, and data extracted in duplicate using predefined criteria. Descriptive analysis will summarize findings, grouped by intervention, with outcomes presented in a narrative synthesis.

**Ethics and Dissemination:** As a scoping review, no ethical approval is required since no primary data collection or direct subject involvement is involved. Findings will be disseminated through peer-reviewed publication, conferences, and relevant digital platforms.

**Registration:** The review protocol has been registered within the Open Science Framework database (https://doi.org/10.17605/OSF.IO/39DSQ)

**Key Messages:**

- **What is already known on this topic** –While several reviews have explored prenatal interventions for gastroschisis, they are often limited in scope— focusing on surgical approaches alone—and frequently lack systematic methodology, limiting the reliability and comprehensiveness of their findings.
- **What this study hopes to add** – This scoping review aims to systematically map and evaluate all published prenatal interventions for gastroschisis, including both surgical and non-surgical approaches.
- **How this study might affect research, practice or policy** – By identifying existing evidence and gaps, this review may inform future research priorities, support clinical decision-making, and contribute to evaluating the potential role of fetal therapy in improving outcomes for gastroschisis.

## INTRODUCTION

Gastroschisis (GS) is one of the most prevalent congenital gastrointestinal malformations, with a rising worldwide incidence of 4-5 per 10,000 live births [1,2]. The condition is characterized by a full-thickness abdominal wall defect, typically to the right of the umbilicus, through which the intestines herniate into the amniotic cavity [3]. The exposed bowel is subject to exposure to amniotic fluid, leading to inflammation, thickening, and subsequent intestinal injury [4,5]. This bowel injury is the primary cause of morbidity and mortality in fetuses with gastroschisis[6].

Although intestinal damage in gastroschisis begins prenatally, current management is focused on postnatal interventions [7,8]. These include neonatal resuscitation, surgical repair of the defect through primary closure or the use of a preformed silo for gradual intestinal reduction and delayed closure, and intensive nutritional management [6,9]. A critical aspect of postnatal care is weaning infants from total parenteral nutrition (TPN) to full enteral feeds as early as possible. Despite these efforts, neonates with gastroschisis often face prolonged hospitalization, increased morbidity related to long-term TPN dependence, and, in severe cases, intestinal failure requiring transplant or leading to mortality [10–12].

The standard prenatal care for gastroschisis is currently limited to fetal surveillance [13], delivery at term unless compromise of fetal well-being, and route of delivery (whether vaginal or cesarean) based on obstetric indication [3]. Given that bowel injury begins in utero, there is growing interest in exploring prenatal interventions aimed at preventing the onset of intestinal damage or mitigating the severity of the injury [14]. These approaches hold the potential to reduce the significant postnatal morbidity and mortality associated with gastroschisis.

Several innovative prenatal therapies have been investigated in both animal models and human studies to address intestinal injury in gastroschisis [15]. These experimental interventions range from elective preterm delivery and trans-amniotic stem cell therapy to amnio-exchange, amnio-infusion, in-utero repair of the defect, prenatal corticosteroid administration, prenatal diuretic administration, placental support, and anti-inflammatory therapies [8,15,16]. Each of these strategies seeks to reduce inflammation or minimize the intestines’ exposure to amniotic fluid. Despite their potential, especially in fetuses with prenatal predictors of complex gastroschisis [14], none of these interventions have been widely adopted in clinical practice, underscoring the need for further research and validation.

This protocol aims to outline the procedures to conduct a systematic scoping review of prenatal interventions to improve outcomes for gastroschisis, to provide a comprehensive overview of the research landscape. This will allow us to identify key gaps in the literature, limitations in current study designs, and areas where more robust evidence is needed. As a scoping review, this approach is particularly suited for exploring the breadth of evidence in this field, to consolidate the existing evidence, guide future research, and inform clinical practice.

## METHODS AND ANALYSIS

The methodology outlined in this systematic review protocol adheres to the Preferred Reporting Items for Systematic Review and Meta-Analysis Protocols (PRISMA-P) guidelines [17]. Appendix 1 includes the PRISMA-P checklist, detailing where each guideline item is addressed within the protocol, when applicable. Any amendments to this protocol will be discussed in the systematic review report, with a description and rationale provided for the amendment.

This scoping review will be conducted in full adherence to the PRISMA-ScR (Preferred Reporting Items for Systematic Reviews and Meta-Analyses Extension for Scoping Reviews) framework [18]. The guidelines will inform every stage of the review process, including protocol development, data extraction, synthesis of results, and reporting, ensuring a methodologically rigorous and transparent approach throughout.

The review protocol has been registered within the Open Science Framework database (https://doi.org/10.17605/OSF.IO/39DSQ).

## Eligibility criteria

Studies will be selected according to the criteria outlined below.

### PICO framework

We will include studies researching the *population* of fetuses diagnosed with gastroschisis, in either human or animal models, as outlined in our search strings (Table 1).

**Table 1.**
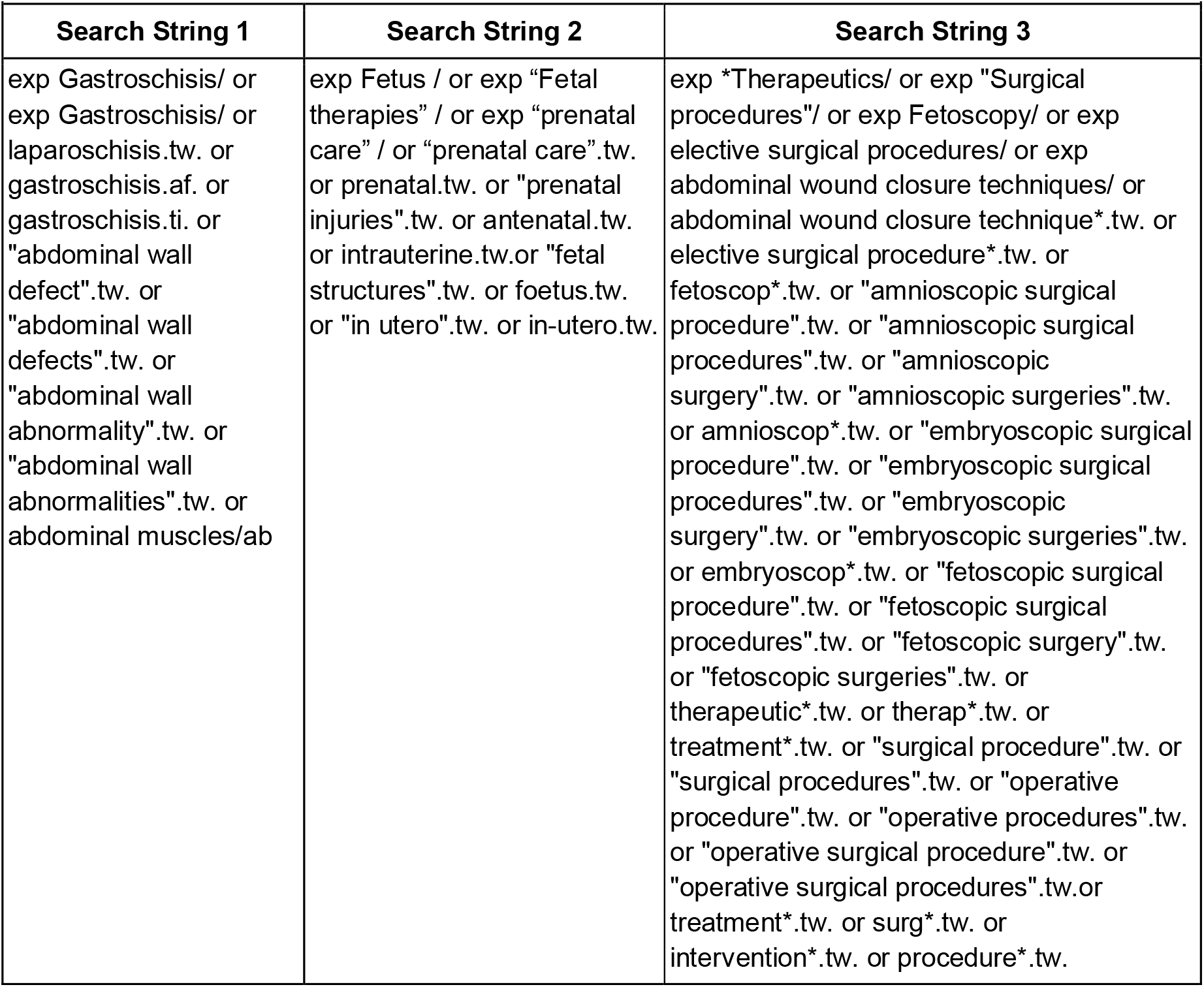
Search strings.

The impact of *prenatal interventions* will be examined. We would consider interventions targeting gastroschisis outcomes, such as elective preterm delivery (EPD), prenatal surgical repair, intraamniotic drug/compound delivery, among others, subject to what exists in the literature.

The *comparison* for animal models will use control data, such as no-intervention groups, while for human studies, it will be made against the standard of care. The standard care for gastroschisis includes prenatal ultrasound surveillance, delivery at term unless compromise of fetal well-being, and route of delivery (whether vaginal or cesarean) based on obstetric indication. Once the infant is born, the care includes postnatal resuscitation, and either primary closure of the defect or gradual intestinal reduction using a preformed silo for delayed closure. Nutritional management is a critical component of the postnatal care plan, focusing on weaning children from total parenteral nutrition (TPN) and transitioning to full enteral feeds, whenever possible [9,19].

#### Outcome measures

Due to the extensive scope of the interventions under consideration, several *outcomes* will be relevant for our study. Of interest are interventions addressing aspects of morbidity or mortality in gastroschisis.

For human studies, we will include a variety of relevant clinical endpoints, with a focus on outcomes related to gastroschisis complications and treatment effectiveness. Primary outcomes will include Days of total parenteral nutrition (TPN) and Length Of Hospital Stay (LOS). If days of TPN are not reported, we will record a related variable, such as Days to full enteral feeds, or Days to start enteral feeds, when available. Secondary outcomes will include prenatal or postnatal death, Complex gastroschisis (defined as atresia, perforation, necrosis, vanishing gastroschisis, short bowel syndrome, or volvulus), and Harms or adverse effects (including but not limited to blood culture-proven sepsis, necrotizing enterocolitis (NEC), wound infection, bowel injury, abdominal compartment syndrome, and short bowel syndrome).

For animal studies, we will focus on outcomes related to intestinal health and intervention efficacy. Primary outcomes will include Intestinal inflammation or Bowel wall thickness as a surrogate measure. Secondary Outcomes will focus on Harms or adverse effects of interventions, and Animal demise.

### Inclusion and exclusion criteria

Studies conducted to investigate prenatal therapies in either animal models or human fetuses with gastroschisis will be included in this review. This will encompass full-text articles in languages other than English and Spanish, which will be translated for review.

The exclusion criteria have been carefully defined to ensure that only studies relevant to this review are included. Studies lacking a control (no-intervention) group will be excluded, as they do not provide a comparative baseline for assessing the effects of the intervention. Case report studies will also be excluded. Furthermore, studies that do not assess outcomes in gastroschisis will be excluded, as they do not provide data relevant to the condition being investigated.

Dissertations, book chapters, letters to the editor, correspondence, and editorials will be excluded from all studied interventions.

For Amnioinfusion (AIF) and Amnioexchange (AE) interventions, studies that fail to perform a separate analysis for patients with oligohydramnios versus those with normal amniotic fluid volume will be also excluded, as these conditions can have distinct impacts on the outcomes.

Studies on preterm delivery for gastroschisis are particularly heterogeneous and often contain potential confounders that can introduce significant biases [20]. Therefore, we applied specific exclusion criteria for this intervention, to focus on what we believe is the most relevant comparison: sie excluded, regardless of the terminology used by the authors. This exclusion criterion also applies to studies that fail to differentiate EPD in their outcome analysis. Additionally, we excluded studies where EPD was performed exclusively in gastroschisis fetuses with compromised well-being or signs of bowel complications during prenatal surveillance. Since EPD in these cases was specifically triggered by fetal compromise, the outcomes of this subgroup may not be representative of the broader gastroschisis population. Lastly, animal studies will be excluded from this intervention, as sufficient evidence is expected from human studies, which provide more clinically relevant data for assessing the efficacy and safety of the intervention.

### Search strategy and sources

Our search strategy was developed using the evidence-based recommendations stated in the PRESS Peer Review of Electronic Search Strategies: 2015 Guideline Statement [21].

We will search MEDLINE (OVID interface), EMBASE (Elsevier interface), Scopus (Elsevier interface), Web of Science (Clarivate Analytics), LILACS Plus Collection (via BIREME/PAHO/WHO), and the Cochrane Library (via Cochrane). Our draft search strategy is included in Appendix 2.

Gray literature will be included to ensure literature saturation and to prevent publication bias. Additional searched sources will be ProQuest Dissertations & Theses Citation Index (Web of Science/Clarivate), ClinicalTrials.gov (NLM), WHO International Clinical Trials Registry (WHO), and ISRCTN registry (Springer Nature). Previous reviews, meta-analyses, and reference lists of studies will also be searched for further studies suitable for inclusion.

No study design or language limits will be imposed on the search. Each database will be searched from the date of inception. Authors of the identified grey literature will be contacted to obtain full reports of data and findings, where available.

The search was optimized by testing the sensitivity and specificity against a set of known relevant articles, peer-reviewed by a librarian experienced in systematic review searching, and further peer-reviewed by a second librarian, not otherwise associated with the project.

The final search strategy employs a combination of controlled vocabulary, keywords, syntax elements such as truncation and wildcards, and synonyms tailored to our PICO framework.

### Study screening and selection

References identified through the electronic search will be uploaded to Covidence (Covidence systematic review software, Veritas Health Innovation, Melbourne, Australia. Available at www.covidence.org) where duplicates will be removed. Two reviewers will independently screen the titles, the abstracts and the full-texts yielded by the search against the inclusion and exclusion criteria. Reviewers will conduct a calibration exercise by screening an initial set of 100 abstracts and full texts, discussing results, and refining screening criteria as needed to ensure consistency. Differences will be resolved through discussion. Once a high level of agreement (>90%) is reached, screening will proceed independently. Any disagreements flagged by Covidence will be resolved through discussion, with a third reviewer consulted to make a final decision if needed.

Records identified, included, and excluded, and the reasons for exclusions will be presented in the PRISMA 2020 Flow Diagram [22].

### Data management

Data will be extracted independently in duplicate and entered into a predetermined data collection table containing the summary of characteristics of included studies and key information. The two databases will be compared, and reviewers will resolve disagreements by discussion to reach consensus.

Calibration exercises will be done for the data extraction phase as well. The two independent reviewers will chart 25 full-text studies and assess interrater agreement through roundtable discussion. Independent charting will begin once a high level of agreement (>80%) is achieved.

Data will be collected and grouped by prenatal intervention of study. Data collection will capture details on study design/characteristics, including sample size and subjects of study (human study, animal study and species), main demographics, intervention details including rationale, gestational age at intervention, year of publication, and outcomes of study. Where possible, outcomes will be expressed in standardized metrics to facilitate data interpretation. When data transformation is deemed the most sensible approach, we will also present the original reported data to ensure transparency and preserve context.

### Critical Appraisal of Individual Sources of Evidence

While a formal risk of bias assessment or quality appraisal of included studies will not be part of this scoping review, any evident methodological weaknesses (e.g., lack of control groups, unclear outcome definitions) will be considered when drawing conclusions.

### Data synthesis

Studies will be grouped by intervention for data synthesis, with outcomes analyzed within each group. Descriptive statistics will be used to summarize the characteristics and findings of the included studies. Information will be presented in tables accompanied by a systematic narrative synthesis of each fetal intervention investigated for gastroschisis. The Synthesis Without Meta-analysis (SWiM) in systematic reviews reporting guideline [23] will be followed to report the findings, with a focus on the appropriate methods for a systematic scoping review.

Clinical and methodological heterogeneity, such as variations in participant characteristics (e.g., gestational age at intervention), outcomes, study design, and co-interventions, will be assessed qualitatively.

### Patient and Public Involvement

Patient and Public Involvement (PPI) was considered during the planning of this scoping review. However, it was deemed unfeasible at this stage, as the most relevant stakeholders— parents of neonates with gastroschisis—are often engaged during clinical care or shortly after birth, limiting their availability for involvement in protocol development. We acknowledge the value of PPI in research and will explore opportunities to incorporate patient and public perspectives in future phases of this ongoing work on gastroschisis.

## DISCUSSION

This protocol outlines what we believe to be the first scoping review focused on prenatal interventions for gastroschisis conducted with a systematic methodology [8,14,15]. The significance of this effort is underscored by the increasing incidence of gastroschisis [1,2], a trend that remains poorly understood but is likely associated with major environmental shifts, given that many known risk factors for the condition—such as tobacco, alcohol, illicit drugs, and socioeconomic disadvantages— are essentially environmental [24,25].

Current postnatal care practices may be insufficient to address the inflammatory processes that begin prenatally [14]. Therefore, investigating fetal interventions could provide an opportunity to improve clinical outcomes. Despite the potential promise of various experimental approaches, none of these interventions have yet been broadly integrated into standard clinical practice [8,14]. This gap highlights the pressing need for further research and validation of these prenatal strategies. With the rapid advancements in fetal therapy and medical technology, it is crucial to reassess therapies that may offer enhanced outcomes in gastroschisis.

Through this scoping review, we aim to map the existing body of literature on prenatal interventions in a systematic way and provide a comprehensive overview that can identify knowledge gaps and guide future research. However, we expect to encounter some limitations. Variability in intervention protocols across different studies can pose a significant challenge. Interventions may differ in terms of timing, dosage, delivery methods, and other procedural factors, resulting in a high degree of heterogeneity in the data. This variation can impair the comparison of study outcomes and hinder the ability to synthesize the evidence. Consequently, the generalizability of findings may be restricted, as the effectiveness of an intervention could be influenced by specific contextual factors unique to each study. Additionally, some interventions, like elective preterm delivery, may be well-researched, while others may lack sufficient investigation. While this imbalance may skew the overall perspective, we view it as an opportunity to highlight areas that warrant further research efforts to improve outcomes in gastroschisis.

## ETHICS AND DISSEMINATION

As this study is a systematic scoping review of previous studies, it does not involve primary data collection or direct contact with subjects. Thus, ethical approval is not required. However, this review will be conducted in accordance with the principles outlined in the PRISMA-ScR (Preferred Reporting Items for Systematic Reviews and Meta-Analyses extension for Scoping Reviews) guidelines to ensure methodological rigor and transparency. By identifying the current landscape of research and highlighting existing gaps, this review will guide future research priorities and inform clinical practice.

The findings will be disseminated through publication in a peer-reviewed journal and presented at relevant conferences in the fields of maternal-fetal medicine, pediatric surgery, or neonatology. Additionally, we will explore opportunities to share the results via academic networks and digital platforms, ensuring that the research community, clinicians, and policymakers can access and benefit from the insights gained.

## Supporting information

PRISMA-P checklist

Search strategy

## Data Availability

https://doi.org/10.17605/OSF.IO/39DSQ

## AUTHORS’ CONTRIBUTIONS

MFV, JLP, MO, EL, SK: Conceived the idea for the study

MFV: Designed and drafted the protocol, and is the guarantor of the review.

JLP, MO, EL, SK: Assisted with the conceptualization of the study and contributed to drafting and revising the protocol.

All authors: Provided input on the study design and protocol development, and contributed to the final manuscript.

## FUNDING

This research received no specific grant from any funding agency in the public, commercial, or not-for-profit sectors.

## COMPETING INTERESTS

The authors declare that they have no competing interests

## ACKNOWLEDGEMENTS

The authors thank the library services at the Children’s Hospital of Philadelphia for their help with the literature search.

Author Varela MF acknowledges the support from the Global PaedSurg Research Training Fellowship for promoting the transmission of crucial research skills across Global PaedSurg collaborators, which greatly contributed to the development of this manuscript.

## Notes

### Competing Interest Statement

The authors have declared no competing interest.

### Funding Statement

This study did not receive any funding

### Author Declarations

As a systematic scoping review, this study will use openly available human data that were originally located at Medline, Enbase, web of science, Scopus, Cochrane Library, ClinicalTrials.gov (NLM), International Clinical Trials Registry Platform (WHO), ISRCTN Registry (Springer Nature), and ProQuest Dissertations & Theses Citation Index (Web of Science/Clarivate).

