## Supplementary material for "Prenatal therapies to improve outcomes in gastroschisis: A systematic scoping review protocol": Search strategy

**Database / Study Registry (including vendor/platform): MEDLINE (Ovid)**

| **Search string** | **Results** |
| --- | --- |
| 1. exp Gastroschisis/ or exp Gastroschisis/ or laparoschisis.tw. or gastroschisis.af. or gastroschisis.ti. or "abdominal wall defect".tw. or "abdominal wall defects".tw. or "abdominal wall abnormality".tw. or "abdominal wall abnormalities".tw. or abdominal muscles/ab | 5,548 |
| 2. exp Fetus / or exp “Fetal therapies” / or exp “prenatal care” / or “prenatal care”.tw. or prenatal.tw. or "prenatal injuries".tw. or antenatal.tw. or intrauterine.tw.or "fetal structures".tw. or foetus.tw. or "in utero".tw. or in-utero.tw. | 402,333 |
| 3. exp *Therapeutics/ or exp "Surgical procedures"/ or exp Fetoscopy/ or exp elective surgical procedures/ or exp abdominal wound closure techniques/ or abdominal wound closure technique*.tw. or elective surgical procedure*.tw. or fetoscop*.tw. or "amnioscopic surgical procedure".tw. or "amnioscopic surgical procedures".tw. or "amnioscopic surgery".tw. or "amnioscopic surgeries".tw. or amnioscop*.tw. or "embryoscopic surgical procedure".tw. or "embryoscopic surgical procedures".tw. or "embryoscopic surgery".tw. or "embryoscopic surgeries".tw. or embryoscop*.tw. or "fetoscopic surgical procedure".tw. or "fetoscopic surgical procedures".tw. or "fetoscopic surgery".tw. or "fetoscopic surgeries".tw. or therapeutic*.tw. or therap*.tw. or treatment*.tw. or "surgical procedure".tw. or "surgical procedures".tw. or "operative procedure".tw. or "operative procedures".tw. or "operative surgical procedure".tw. or "operative surgical procedures".tw.or treatment*.tw. or surg*.tw. or intervention*.tw. or procedure*.tw. | 1,324,1125 |
| 4. 1 AND 2 AND 3 | 665 |

**Database / Study Registry (including vendor/platform):** **EMBASE (Elsevier)**

| **Search string** | **Results** |
| --- | --- |
| 1. 'gastroschisis'/exp OR 'laparoschisis':ti,ab,kw OR 'abdominal wall defect':ti,ab,kw OR 'abdominal wall defects':ti,ab,kw OR 'abdominal wall abnormality':ti,ab,kw OR 'abdominal wall abnormalities':ti,ab,kw OR 'abdominal wall musculature'/de | 27,269 |
| 2. 'fetus'/exp OR 'fetal therapy'/exp OR 'prenatal care':ti,ab,kw OR 'prenatal':ti,ab,kw OR 'prenatal injuries':ti,ab,kw OR 'antenatal':ti,ab,kw OR 'intrauterine':ti,ab,kw OR 'fetal structures':ti,ab,kw OR 'foetus':ti,ab,kw OR 'in utero':ti,ab,kw OR 'in-utero':ti,ab,kw | 511,128 |
| 3. 'therapy'/exp/mj OR 'therapy' OR 'fetoscopy'/exp OR 'fetoscopy' OR 'elective surgery'/exp OR 'elective surgery' OR 'abdominal wound closure'/exp OR 'abdominal wound closure' OR 'abdominal wound closure technique*':ti,ab,kw OR 'elective surgical procedure*':ti,ab,kw OR 'fetoscop*':ti,ab,kw OR 'amnioscopic surgical procedure':ti,ab,kw OR 'amnioscopic surgical procedures':ti,ab,kw OR 'amnioscopic surgery':ti,ab,kw OR 'amnioscopic surgeries':ti,ab,kw OR 'amnioscop*':ti,ab,kw OR 'embryoscopic surgical procedure':ti,ab,kw OR 'embryoscopic surgical procedures':ti,ab,kw OR 'embryoscopic surgery':ti,ab,kw OR 'embryoscopic surgeries':ti,ab,kw OR 'embryoscop*':ti,ab,kw OR 'fetoscopic surgical procedure':ti,ab,kw OR 'fetoscopic surgical procedures':ti,ab,kw OR 'fetoscopic surgery':ti,ab,kw OR 'fetoscopic surgeries':ti,ab,kw OR 'therapeutic*':ti,ab,kw OR 'therap*':ti,ab,kw OR 'treatment*':ti,ab,kw OR 'surgical procedure':ti,ab,kw OR 'surgical procedures':ti,ab,kw OR 'operative procedure':ti,ab,kw OR 'operative procedures':ti,ab,kw OR 'operative surgical procedure':ti,ab,kw OR 'operative surgical proceduresor treatment*':ti,ab,kw OR 'surg*':ti,ab,kw OR 'intervention*':ti,ab,kw OR 'procedure*':ti,ab,kw | 19,261,662 |
| 4. 1 AND 2 AND 3 | 1,067 |

**Database / Study Registry (including vendor/platform):** **Scopus (Elsevier)**

| **Search string** | **Results** |
| --- | --- |
| 1. INDEXTERMS (gastroschisis) OR INDEXTERMS ("Abdominal Wall abnormalities") OR TITLE-ABS (laparoschisis) OR ALL (gastroschisis) OR TITLE (gastroschisis) OR TITLE-ABS ("abdominal wall defect") OR TITLE-ABS ("abdominal wall defects") OR TITLE-ABS ("abdominal wall abnormality") OR TITLE-ABS ("abdominal wall abnormalities") OR INDEXTERMS ("abdominal muscles") | 21,158 |
| 2. INDEXTERMS (fetus) OR INDEXTERMS ("Fetal therapies") OR INDEXTERMS ("prenatal care") OR TITLE-ABS ("prenatal care") OR TITLE-ABS (prenatal) OR TITLE-ABS ("prenatal injuries") OR TITLE-ABS (antenatal) OR TITLE-ABS (intrauterine) OR TITLE-ABS ("fetal structures") OR TITLE-ABS (foetus) OR TITLE-ABS ("in utero") OR TITLE-ABS (in-utero) | 631,250 |
| 3. INDEXTERMS ( therapeutics ) OR INDEXTERMS ( "Surgical procedures" ) OR INDEXTERMS ( fetoscopy ) OR INDEXTERMS ( "elective surgical procedures" ) OR INDEXTERMS ( "abdominal wound closure techniques" ) OR TITLE-ABS ( "abdominal wound closure technique*" ) OR TITLE-ABS ( "elective surgical procedure*" ) OR TITLE-ABS ( fetoscop* ) OR TITLE-ABS ( "amnioscopic surgical procedure" ) OR TITLE-ABS ( "amnioscopic surgical procedures" ) OR TITLE-ABS ( "amnioscopic surgery" ) OR TITLE-ABS ( "amnioscopic surgeries" ) OR TITLE-ABS ( amnioscop* ) OR TITLE-ABS ( "embryoscopic surgical procedure" ) OR TITLE-ABS ( "embryoscopic surgical procedures" ) OR TITLE-ABS ( "embryoscopic surgery" ) OR TITLE-ABS ( "embryoscopic surgeries" ) OR TITLE-ABS ( embryoscop* ) OR TITLE-ABS ( "fetoscopic surgical procedure" ) OR TITLE-ABS ( "fetoscopic surgical procedures" ) OR TITLE-ABS ( "fetoscopic surgery" ) OR TITLE-ABS ( "fetoscopic surgeries" ) OR TITLE-ABS ( therapeutic* ) OR TITLE-ABS ( therap* ) OR TITLE-ABS ( treatment* ) OR TITLE-ABS ( "surgical procedure" ) OR TITLE-ABS ( "surgical procedures" ) OR TITLE-ABS ( "operative procedure" ) OR TITLE-ABS ( "operative procedures" ) OR TITLE-ABS ( "operative surgical procedure" ) OR TITLE-ABS ( "operative surgical procedures" ) OR TITLE-ABS ( treatment* ) OR TITLE-ABS ( surg* ) OR TITLE-ABS ( intervention* ) OR TITLE-ABS ( procedure* ) | 16,291,678 |
| 4. 1 AND 2 AND 3 | 1,348 |

**Database / Study Registry (including vendor/platform):** **Web of Science (Clarivate)**

| **Search string** | **Results** |
| --- | --- |
| 1. ALL=Gastroschisis OR ALL=Gastroschisis OR ALL="Abdominal Wall abnormalities" OR (TI=laparoschistis OR AB=laparoschistis) OR ALL=gastroschisis OR TI=gastroschisis OR (TI="abdominal wall defect" OR AB="abdominal wall defect") OR (TI="abdominal wall defects" OR AB="abdominal wall defects") OR (TI="abdominal wall abnormality" OR AB="abdominal wall abnormality") OR (TI="abdominal wall abnormalities" OR AB="abdominal wall abnormalities") OR ALL="abdominal muscles" | 7,654 |
| 2. ALL=Fetus OR ALL=”Fetal therapies” OR ALL=”prenatal care” OR (TI="prenatal care" OR AB="prenatal care") OR (TI=prenatal OR AB=prenatal) OR (TI="prenatal injuries" OR AB="prenatal injuries") OR (TI=antenatal OR AB=antenatal) OR (TI=intrauterine OR AB=intrauterine) OR (TI="fetal structures” OR AB=”fetal structures”) OR (TI=foetus OR AB=foetus) OR (TI="in utero" OR AB="in utero") OR (TI=in-utero OR AB=in-utero) | 322,559 |
| 3. ALL=Therapeutics OR ALL="Surgical procedures" OR ALL=Fetoscopy OR ALL="elective surgical procedures" OR ALL="abdominal wound closure techniques" OR (TI="abdominal wound closure technique*" OR AB="abdominal wound closure technique*") OR (TI="elective surgical procedure*" OR AB="elective surgical procedure*") OR (TI=fetoscop* OR AB=fetoscop*) OR (TI="amnioscopic surgical procedure" OR AB="amnioscopic surgical procedure") OR (TI="amnioscopic surgical procedures" OR AB="amnioscopic surgical procedures") OR (TI="amnioscopic surgery" OR AB="amnioscopic surgery") OR (TI="amnioscopic surgeries" OR AB="amnioscopic surgeries") OR (TI=amnioscop* OR AB=amnioscop*) OR (TI="embryoscopic surgical procedure" OR AB="embryoscopic surgical procedure") OR (TI="embryoscopic surgical procedures" OR AB="embryoscopic surgical procedures") OR (TI="embryoscopic surgery" OR AB="embryoscopic surgery") OR (TI="embryoscopic surgeries" OR AB="embryoscopic surgeries") OR (TI=embryoscop* OR AB=embryoscop*) OR (TI="fetoscopic surgical procedure" OR AB="fetoscopic surgical procedure") OR (TI="fetoscopic surgical procedures" OR AB="fetoscopic surgical procedures") OR (TI="fetoscopic surgery" OR AB="fetoscopic surgery") OR (TI="fetoscopic surgeries" OR AB="fetoscopic surgeries") OR (TI=therapeutic* OR AB=therapeutic*) OR (TI=therap* OR AB=therap*) OR (TI=treatment* OR AB=treatment*) OR (TI="surgical procedure" OR AB="surgical procedure") OR (TI="surgical procedures" OR AB="surgical procedures") OR (TI="operative procedure" OR AB="operative procedure") OR (TI="operative procedures" OR AB="operative procedures") OR (TI="operative surgical procedure" OR AB="operative surgical procedure") OR (TI="operative surgical procedures” OR AB="operative surgical procedures") OR (TI=treatment* OR AB=treatment*) OR (TI=surg* OR AB=surg*) OR (TI=intervention* OR AB=intervention*) OR (TI=procedure* OR AB=procedure*) | 12,046,397 |
| 4. 1 AND 2 AND 3 | 492 |

**Database:** **Cochrane Library (Cochrane)**

| **Search string** | **Results** |
| --- | --- |
| 1. [mh Gastroschisis] OR [mh Gastroschisis] OR [mh "Abdominal Wall abnormalities"] OR laparoschisis:ti,ab OR gastroschisis OR gastroschisis:ti OR "abdominal wall defect":ti,ab OR "abdominal wall defects":ti,ab OR "abdominal wall abnormality":ti,ab OR "abdominal wall abnormalities":ti,ab OR [mh ^"abdominal muscles"] | 942 |
| 2. [mh Fetus] OR [mh “Fetal therapies”] OR [mh “prenatal care”] OR "prenatal care":ti,ab OR prenatal:ti,ab OR "prenatal injuries":ti,ab OR antenatal:ti,ab OR intrauterine OR "fetal structures":ti,ab OR foetus:ti,ab OR "in utero":ti,ab OR in-utero:ti,ab | 26,451 |
| 3. [mh Therapeutics] OR [mh "Surgical procedures"] OR [mh Fetoscopy] OR [mh "elective surgical procedures"] OR [mh "abdominal wound closure techniques"] OR (abdominal wound closure NEXT technique*):ti,ab OR (elective surgical NEXT procedure*):ti,ab OR fetoscop*:ti,ab OR amnioscopic surgical procedure:ti,ab OR amnioscopic surgical procedures:ti,ab OR amnioscopic surgery:ti,ab OR amnioscopic surgeries:ti,ab OR amnioscop*:ti,ab OR embryoscopic surgical procedure:ti,ab OR embryoscopic surgical procedures:ti,ab OR embryoscopic surgery:ti,ab OR embryoscopic surgeries:ti,ab OR embryoscop*:ti,ab OR fetoscopic surgical procedure:ti,ab OR fetoscopic surgical procedures:ti,ab OR fetoscopic surgery:ti,ab OR fetoscopic surgeries:ti,ab OR therapeutic*:ti,ab OR therap*:ti,ab OR treatment*:ti,ab OR surgical procedure:ti,ab OR surgical procedures:ti,ab OR operative procedure:ti,ab OR operative procedures:ti,ab OR operative surgical procedure:ti,ab OR operative surgical procedures:ti,ab OR treatment*:ti,ab OR surg*:ti,ab OR intervention*:ti,ab OR procedure*:ti,ab | 1,650,610 |
| 4. 1 AND 2 AND 3 | 27 |

**Database: LILACS Plus Collection (BIREME/PAHO/WHO)**

**Date: 10/24/24**

| **Search string** | **Results** |
| --- | --- |
| 1. (mh:"Gastroschisis" OR tw:"Gastroschisis" OR tw:"laparoschisis" OR tw:"abdominal wall defect" OR tw:"abdominal wall defects" OR tw:"abdominal wall abnormality" OR tw:"abdominal wall abnormalities" OR mh:"Abdominal Muscles") | 1,427 |
| 2. (mh:"Fetus" OR mh:"Fetal Therapies" OR mh:"Prenatal Care" OR tw:"prenatal care" OR tw:prenatal OR tw:"prenatal injuries" OR tw:antenatal OR tw:intrauterine OR tw:"fetal structures" OR tw:fetus OR tw:"in utero" OR tw:in-utero) | 59,562 |
| 3. (mh:"Therapeutics" OR mh:"Surgical Procedures, Operative" OR mh:"Fetoscopy" OR mh:"Elective Surgical Procedures" OR mh:"Abdominal Wound Closure Techniques" OR tw:"abdominal wound closure technique" OR tw:"elective surgical procedure" OR tw:fetoscop$ OR tw:"amnioscopic surgical procedure" OR tw:"amnioscopic surgery" OR tw:amnioscop$ OR tw:"embryoscopic surgical procedure" OR tw:"embryoscopic surgery" OR tw:embryoscop$ OR tw:"fetoscopic surgical procedure" OR tw:"fetoscopic surgery" OR tw:therapeut$ OR tw:therap$ OR tw:treatment$ OR tw:"surgical procedure" OR tw:"operative procedure" OR tw:surg$ OR tw:intervent$ OR tw:procedure$) | 1,108,639 |
| 4. 1 AND 2 AND 3 | 149 |

**Database: ClinicalTrials.gov (NLM)**

| **Search string** | **Results** |
| --- | --- |
| (gastroschisis OR laparoschisis OR "abdominal wall defect" OR "abdominal wall abnormality") AND (fetus OR fetal OR prenatal OR antenatal OR intrauterine OR "in utero") AND (therapy OR treatment OR surgery OR surgical OR fetoscopy OR amnioscopy OR embryoscopy OR intervention OR procedure) | **13** |

**Database: International Clinical Trials Registry Platform (WHO)**

| **Search string** | **Results** |
| --- | --- |
| (gastroschisis OR laparoschisis OR "abdominal wall defect" OR "abdominal wall abnormality") AND (fetus OR fetal OR prenatal OR antenatal OR intrauterine OR "in utero") AND (therapy OR treatment OR surgery OR surgical OR fetoscopy OR amnioscopy OR embryoscopy OR intervention OR procedure) | **1** |

**Database: ISRCTN Registry (Springer Nature)**

| **Search string** | **Results** |
| --- | --- |
| Text search: fetus OR fetal OR prenatal OR antenatal OR "in utero"  Condition: gastroschisis OR laparoschisis OR "abdominal wall defect" OR "abdominal wall abnormality"  Interventions: therapy OR treatment OR surgery OR surgical OR fetoscopy OR amnioscopy OR embryoscopy OR intervention OR procedure | 781 |

**Database / Study Registry (including vendor/platform):** **ProQuest Dissertations & Theses Citation Index (Web of Science/Clarivate)**

| **Search string** | **Results** |
| --- | --- |
| 1. ALL=Gastroschisis OR ALL=Gastroschisis OR ALL="Abdominal Wall abnormalities" OR (TI=laparoschistis OR AB=laparoschistis) OR ALL=gastroschisis OR TI=gastroschisis OR (TI="abdominal wall defect" OR AB="abdominal wall defect") OR (TI="abdominal wall defects" OR AB="abdominal wall defects") OR (TI="abdominal wall abnormality" OR AB="abdominal wall abnormality") OR (TI="abdominal wall abnormalities" OR AB="abdominal wall abnormalities") OR ALL="abdominal muscles" | 232 |
| 2. ALL=Fetus OR ALL=”Fetal therapies” OR ALL=”prenatal care” OR (TI="prenatal care" OR AB="prenatal care") OR (TI=prenatal OR AB=prenatal) OR (TI="prenatal injuries" OR AB="prenatal injuries") OR (TI=antenatal OR AB=antenatal) OR (TI=intrauterine OR AB=intrauterine) OR (TI="fetal structures” OR AB=”fetal structures”) OR (TI=foetus OR AB=foetus) OR (TI="in utero" OR AB="in utero") OR (TI=in-utero OR AB=in-utero) | 18.150 |
| 3. ALL=Therapeutics OR ALL="Surgical procedures" OR ALL=Fetoscopy OR ALL="elective surgical procedures" OR ALL="abdominal wound closure techniques" OR (TI="abdominal wound closure technique*" OR AB="abdominal wound closure technique*") OR (TI="elective surgical procedure*" OR AB="elective surgical procedure*") OR (TI=fetoscop* OR AB=fetoscop*) OR (TI="amnioscopic surgical procedure" OR AB="amnioscopic surgical procedure") OR (TI="amnioscopic surgical procedures" OR AB="amnioscopic surgical procedures") OR (TI="amnioscopic surgery" OR AB="amnioscopic surgery") OR (TI="amnioscopic surgeries" OR AB="amnioscopic surgeries") OR (TI=amnioscop* OR AB=amnioscop*) OR (TI="embryoscopic surgical procedure" OR AB="embryoscopic surgical procedure") OR (TI="embryoscopic surgical procedures" OR AB="embryoscopic surgical procedures") OR (TI="embryoscopic surgery" OR AB="embryoscopic surgery") OR (TI="embryoscopic surgeries" OR AB="embryoscopic surgeries") OR (TI=embryoscop* OR AB=embryoscop*) OR (TI="fetoscopic surgical procedure" OR AB="fetoscopic surgical procedure") OR (TI="fetoscopic surgical procedures" OR AB="fetoscopic surgical procedures") OR (TI="fetoscopic surgery" OR AB="fetoscopic surgery") OR (TI="fetoscopic surgeries" OR AB="fetoscopic surgeries") OR (TI=therapeutic* OR AB=therapeutic*) OR (TI=therap* OR AB=therap*) OR (TI=treatment* OR AB=treatment*) OR (TI="surgical procedure" OR AB="surgical procedure") OR (TI="surgical procedures" OR AB="surgical procedures") OR (TI="operative procedure" OR AB="operative procedure") OR (TI="operative procedures" OR AB="operative procedures") OR (TI="operative surgical procedure" OR AB="operative surgical procedure") OR (TI="operative surgical procedures” OR AB="operative surgical procedures") OR (TI=treatment* OR AB=treatment*) OR (TI=surg* OR AB=surg*) OR (TI=intervention* OR AB=intervention*) OR (TI=procedure* OR AB=procedure*) | 954,988 |
| 4. 1 AND 2 AND 3 | 12 |
